# Pathway-Specific Polygenic Risk Scores for Blood Pressure Traits in a West African Cohort

**DOI:** 10.1101/2025.06.30.25330577

**Authors:** Gregory Bormes, Vanessa Robbin, Tinashe Chikwore, Yuji Zhang, Ananyo Choudhury, Scott Hazelhurst, Neil A. Hanchard, Sally N. Adebamowo, Adebowale A. Adeyemo, Bamidele Tayo

## Abstract

**Introduction:** Genome-wide polygenic risk scores (PRS) are useful for stratifying individuals’ risk for polygenic diseases such as hypertension. However, a downside of genome-wide PRS is the lack of information about the distribution of risk burden across biologic pathways. We used pathway-specific PRS to investigate these effects within common anti-hypertensive therapy-target pathways on disease risk in a cohort of West Africans.

**Methods:** A total of 11 pathways comprising 1,149 unique genes were selected based on the targets of common anti-hypertensive agents. Pathway-specific PRS for hypertension (individuals with systolic blood pressure (SBP) ≥140 mmHg, diastolic blood pressure (DBP) ≥90 mmHg, or taking anti-hypertensive medications) were computed in a cohort of 2,395 individuals. The model was then validated and tested in 1,614 and 966 separate individuals, respectively. All participants were recruited from the International Collaborative Study on Hypertension in Blacks.

**Results:** In combined pathways analysis, PRS predicted risk better than base models fitted with only sex, age, and principal components. Compared to base models without the PRS, the incremental increases in R^2^ attributable to inclusion of PRS in predictive models were 2.6% for SBP (p = 0.009); 1.4% for DBP (p = 0.012); and 1.1% for mean arterial pressure (MAP) (p = 0.044.) PRS from certain pathways (MAPK, cAMP, and adrenergic signaling in cardiomyocytes) could stratify individuals in the top and bottom deciles for DBP. Adrenergic signaling in cardiomyocytes was also predictive of MAP when comparing top and bottom deciles.

**Conclusions:** Combined pathway polygenic risk scores constructed from genes in well-defined genetic pathways predict hypertension risk in individuals of African ancestry. However, pathway-specific PRS’s relatively low predictability supports the need to explore the broader influence of genetic, environmental, and epigenetic factors that cannot be captured by pathway-specific PRS alone.

## Introduction

The growing prevalence of hypertension across populations is a global health concern with an estimated 1.28 billion individuals currently impacted, with fewer than one-quarter of those considered controlled.^1,2^ Hypertension carries a significant mortality risk and uncontrolled hypertension is associated with significant sequelae, including stroke and myocardial infarction. Despite the long history of antihypertensive therapy, affected individuals frequently respond unpredictably and/or ineffectively to its treatment. ^2,3^ Understanding the genetic contributions to the progression of this multifactorial disease has been an important area of research in cardiovascular disease.^2,3^ Improving our understanding of the underpinnings of hypertension pathophysiology may better allow clinicians to identify patients with high risk for closer monitoring and suggest the use of more targeted therapies.

Genome-wide polygenic risk scores (PRS) are a promising tool for stratifying individuals’ risk for disease by aggregating the effects of hundreds or thousands of genes which often span the entire genome.^4^ While the traditional genome-wide approach has the potential benefit of better characterizing widely polygenic traits by identifying unexpected contributors to certain traits, there are some associated drawbacks. For traits and disorders that involve dysregulation across multiple systems and pathways, such as hypertension, a key limitation of PRS is that they offer no insight into how overall genetic risk is apportioned among distinct biological pathways. This can limit the understanding of the underlying mechanisms of disease and thus limit interpretability of the PRS. Narrowing the scope of PRS analyses to putative biological pathways can offer more specific insights into disease processes, individual risk, and medication efficacy.^3,5^ Accordingly, there has been strong interest in pathway-specific polygenic risk scores in recent years in a wide variety of conditions, including coronary artery disease, Alzheimer’s, and Parkinson’s disease.^5–8^ Notably, pathway-specific PRS have been shown to outperform conventional PRS in stratifying individuals by their disease risk.^9–11^

Many of the most commonly-used anti-hypertensive medications act on specific, well-defined biological pathways.^2^ Previous pathway-based PRS studies in European cohorts have identified several genetic pathways implicated in hypertension, but with limited generalizability to other populations.^3^ For example, a PRS for hypertension was shown to explain 8.0% of systolic blood pressure and 7.8% of diastolic blood pressure variability in white individuals vs. 3.5% and 3.1% in black individuals, respectively.^4^ Given how poorly existing PRS models perform in African ancestry populations,^4,12^ we investigated how genetic variation within genes constituting common anti-hypertensive medication target-pathways affect hypertension risk in a cohort of Africans residing in Nigeria. We calculated pathway-specific PRS for hypertension using three separate cohorts from the same homogeneous ancestry group to create, validate, and test the model. Pathways of interest were selected based on targets of common first-line antihypertensive medications^3^ (Table 1).

**Table 1.** Target Pathways of Common Anti-Hypertensive Medications.

| Pathway | KEGG ID | Number of Genes |
| --- | --- | --- |
| Adrenergic Signaling in Cardiomyocytes<sup>b,c,d</sup> | hsa04261 | 170 |
| Aldosterone Synthesis and Secretion<sup>b</sup> | hsa04925 | 116 |
| Calcium Signaling<sup>b,c,d</sup> | hsa04020 | 269 |
| Cardiac Muscle Contraction<sup>a,c,d</sup> | hsa04260 | 93 |
| MAPK Signaling Pathway<sup>c</sup> | hsa04010 | 343 |
| Neuroactive Ligand Receptor Interaction<sup>b,d</sup> | hsa04080 | 392 |
| Renin-Angiotensin System<sup>a,b</sup> | hsa04614 | 24 |
| Renin Secretion<sup>b</sup> | hsa04924 | 79 |
| cGMP-PKG Signaling Pathway<sup>d</sup> | hsa04022 | 183 |
| cAMP Signaling Pathway<sup>d</sup> | hsa04024 | 245 |
| Vascular Smooth Muscle Contraction<sup>b,c</sup> | hsa04270 | 146 |

a. ACE Inhibitor Target; b. Angiotensin Receptor Blocker Target; c. Calcium Channel Blocker Target; d. Beta-Blocker Target

## Methods

### Study participants

Data analyzed in this study were from three non-overlapping Nigerian cohorts from the same homogeneous ancestry group whose sampling frame was provided by the International Collaborative Study on Hypertension in Blacks (ICSHIB), as described in detail elsewhere.^13,14^ The base cohort consists of over 15,000 participants recruited from the Yoruba-speaking city of Ibadan and the town of Igbo-Ora in Oyo State, Nigeria. The project was reviewed and approved by Loyola University Chicago and the University of Ibadan. All participants signed informed consent administered in either English or Yoruba language.

### Phenotype measurement

A screening exam was performed by trained research staff using a standardized protocol. ^15^ Information on family and medical history was obtained from each participant. Blood pressure (BP) measurements were conducted by staff trained and certified by a previously described procedure.^13,15^ An oscillometric device which had previously been evaluated in similar field settings was used for all BP measurements.^15^ Three measurements were taken three minutes apart and the average of the final two was used in the analysis. Individuals with systolic blood pressure (SBP) ≥140 mmHg, diastolic blood pressure (DBP) ≥90 mmHg or on anti-hypertensive medication at time of exam were defined as hypertensive. Participants identified with hypertension by these criteria were referred for treatment and follow-up by a physician after detection at the screening exam.

### Genotyping and imputation quality assessment

SNP array data from the three cohorts underwent quality control measures as described previously by Nandakumar et al.^15,16^ The genotyping for the cohorts was carried out with the Illumina Human Exome BeadChip v1.0 (NEX1KG cohort, n=2395), Affymetrix Genome-Wide Human SNP Array 6.0 (NGR1KG cohort, n=1614), and Affymetrix Axiom Genome-Wide KP UCSF AFR Array (NAX1KG cohort, n=966). Using the Sanger Imputation Service,^17^ we performed genotype imputation separately for each cohort to infer missing genotypes or genotypes at ungenotyped loci by using the 1000 Genomes Phase 3 reference panel.^18^ Imputed variants with imputation quality value less than 0.3 or with minor allele frequency less than 0.05 were filtered out as part of post imputation quality control procedures.

### Pathway selection and SNP mapping

Biological pathways were selected based primarily on their known involvement in anti-hypertensive medication mechanisms. The KEGG Pathway Database was used to identify and select pathways targeted by common first-line anti-hypertensive medications. Genetic pathway targets of beta-blockers, calcium channel blockers, ACE inhibitors, and angiotensin receptor blockers were included (Table 1). The selected pathways represent key processes in blood pressure pathophysiology, regulation, and therapeutic response.

For each pathway, we extracted single nucleotide polymorphisms (SNPs) available in our imputed GWAS datasets by mapping SNPs to pathway genes based on coordinates from the Genome Reference Consortium Human Build 37.

### Cohort specification

Three independent cohorts served as discovery, validation, and target cohorts.^13,14^ The discovery cohort (NEX1KG, n = 2,295) was used for initial SNP-BP associations testing and effect size estimation. The validation cohort (NGR1KG, n = 1,614) was used for PRS optimization and threshold selection. The target cohort (NAX1KG, n = 996) was used to assess the performance of the PRS model.

### PRS calculation and analysis

#### Discovery cohort

Polygenic Risk Scores were calculated for SBP, DBP, mean arterial pressure (MAP), and pulse pressure (PP) for each of the eleven target pathways. Additionally, using genes aggregated from all pathways, combined-pathway PRS was calculated for each trait.

Analyses of SNP-BP associations in the discovery cohort were performed for BP as a continuous trait by fitting additive genetic model for each SNP in a linear regression model adjusted for age and sex. The first five genetic principal components were also included as covariates to correct for effects of any potential population stratification.^19^ For each SNP, the risk allele was defined as the trait-increasing allele, and every SNP was coded as the count or dosage of the respective risk allele per genotype. Adjusted BP measures were used for individuals on anti-hypertensive medications by adding 10 and 15 mmHg to measured diastolic BP and systolic BP, respectively, according to a standard method used in BP GWAS.^20,21^ Training of the PRS model was performed on the discovery cohort using a “clumping and thresholding” method.

To address linkage disequilibrium within the cohort, clumping was performed using the software PLINK^22^ ^23^ version 1.9. SNPs within 250kb of the index SNP were considered for clumping. SNPs with r^2^ >0.1 based on maximum likelihood haplotype frequency estimates were considered to be in linkage disequilibrium with the index SNP and were thus excluded from analysis.

PRS were calculated for combined pathways at seven p-value thresholds (0.0010, 0.0025, 0.0050, 0.0075, 0.0100, 0.0250, 0.0500) and corresponding effect sizes for the SNPs were calculated and extracted for use as weights in the validation and target cohorts. In addition, pathway-specific PRSs were calculated at each threshold. We used the default settings for PRS calculation in PLINK. ^23^

#### Validation cohort

We used the validation cohort to identify the best fitting PRS model by fitting linear regression model of BP traits on PRS at the 7 different *p-value* thresholds with sex, age, and the top five principal components included as covariates.^16^ The best-fit *p-value* threshold was chosen to maximize the incremental R², calculated as R²_increment_ = R²_full_ - R²_base_, where R²_full_ includes PRS and covariates, while R²_base_ includes only covariates.

#### Target cohort

To assess prediction performance of the selected PRS model for respective BP traits in both individual and combined pathways, we applied the PRS model to an independent target cohort. Performance was evaluated based on the incremental R² similarly to the approach used in the validation cohort. Samples in the target cohort were divided into deciles according to the PRS values. The mean and SD of each decile were calculated for each BP trait. To determine if PRS could reliably differentiate among mean BP traits, the first and tenth deciles were compared using a two-sample t-test.

### Quality control and software

All analyses were performed using R version 4.3.3 for statistical computing and PLINK v1.9 ^23^ for genetic analyses. Quality control measures included checking for genotyped SNP missingness, minor allele frequency, and Hardy-Weinberg equilibrium. Intermediate results, including SNP lists, clumping results, combined pathways and pathway-specific PRS distributions, were documented and archived for reproducibility. Phenotype data were adjusted for covariates including age, sex, and the top five principal components to correct for potential confounding factors and/or population stratification.

## Results

### Cohort Characteristics

The descriptive characteristics of individuals in the discovery, validation and target cohorts are presented in Table 2. There were 2,295 individuals with mean age 48.7±11.2 years, mean SBP of 159.7±29.5 mmHg, and mean DBP of 101.3±18.2 mmHg in the discovery cohort.

**Table 2.** Study Population Characteristics.

| **Variable** | **Discovery Cohort (NEX1KG)** |  | **Target Cohort (NGR1KG)** |  | **Validation Cohort (NAX1KG)** |  |
| --- | --- | --- | --- | --- | --- | --- |
|  | **N** | **Mean (SD)** | **N** | **Mean (SD)** | **N** | **Mean (SD)** |
| Age (years) | 2295 | 48.7 (11.2) | 1614 | 49.0 (14.5) | 966 | 53.1 (11.5) |
| Female | 1683 | 73.3 | 940 | 58.2 | 734 | 76 |
| Male | 612 | 26.7 | 674 | 41.8 | 232 | 24 |
| SBP (mmHg) | 2295 | 159.7 (29.5) | 1614 | 135.4 (29.9) | 966 | 152.0 (31.5) |
| DBP (mmHg) | 2295 | 101.3 (18.2) | 1614 | 83.7 (18.6) | 966 | 93.6 (19.3) |
| PP (mmHg) | 2295 | 58.4 (17.0) | 1614 | 51.7 (16.4) | 966 | 58.4 (17.1) |
| MAP (mmHg) | 2295 | 120.7 (21.1) | 1614 | 100.9 (21.7) | 966 | 113.1 (22.6) |

SBP, systolic blood pressure; DBP, diastolic blood pressure; PP, pulse pressure; MAP, mean arterial pressure.

The validation cohort contained 1,614 individuals with a mean age of 49.0±14.5 years, mean SBP of 135.4±29.9 mmHg, and mean DBP of 83.7±18.6 mmHg. Similarly, the target cohort contained 966 individuals with a mean age of 53.1±11.5 years, mean SBP of 152.0±31.5 mmHg, and mean DBP of 93.6±19.3 mmHg.

### Combined Pathways PRS

Compiling genes across all 11 pathways yielded a list of 1,149 unique genes with a total of 13,237 SNPs present across discovery, validation, and targe cohorts. This gene set was used to calculate the combined pathways PRS for SBP, DBP, MAP, and PP.

After clumping, 1,154 independent SNPs remained for SBP PRS calculation. For SBP, the optimal *p-value* threshold was found to be 0.05, yielding a list of 101 SNPs for analysis, corresponding to 85 unique genes. In our target cohort, the incremental increase in R^2^ due to PRS for SBP was 0.0056 (2.6%, p = 0.009).

When comparing the first and last deciles of PRS scores for SBP, the combined pathways PRS distinguished between SBP with a moderate effect size (t = −3.38, p = 0.00087, Cohen’s d = 0.49). Mean SBP for the first PRS decile was 141.2 mmHg (SD = 33.5); mean SBP for the last PRS decile was 157.0 mmHg (SD = 31.7) (Figures 1a and 2a).

**Figure 1a.**
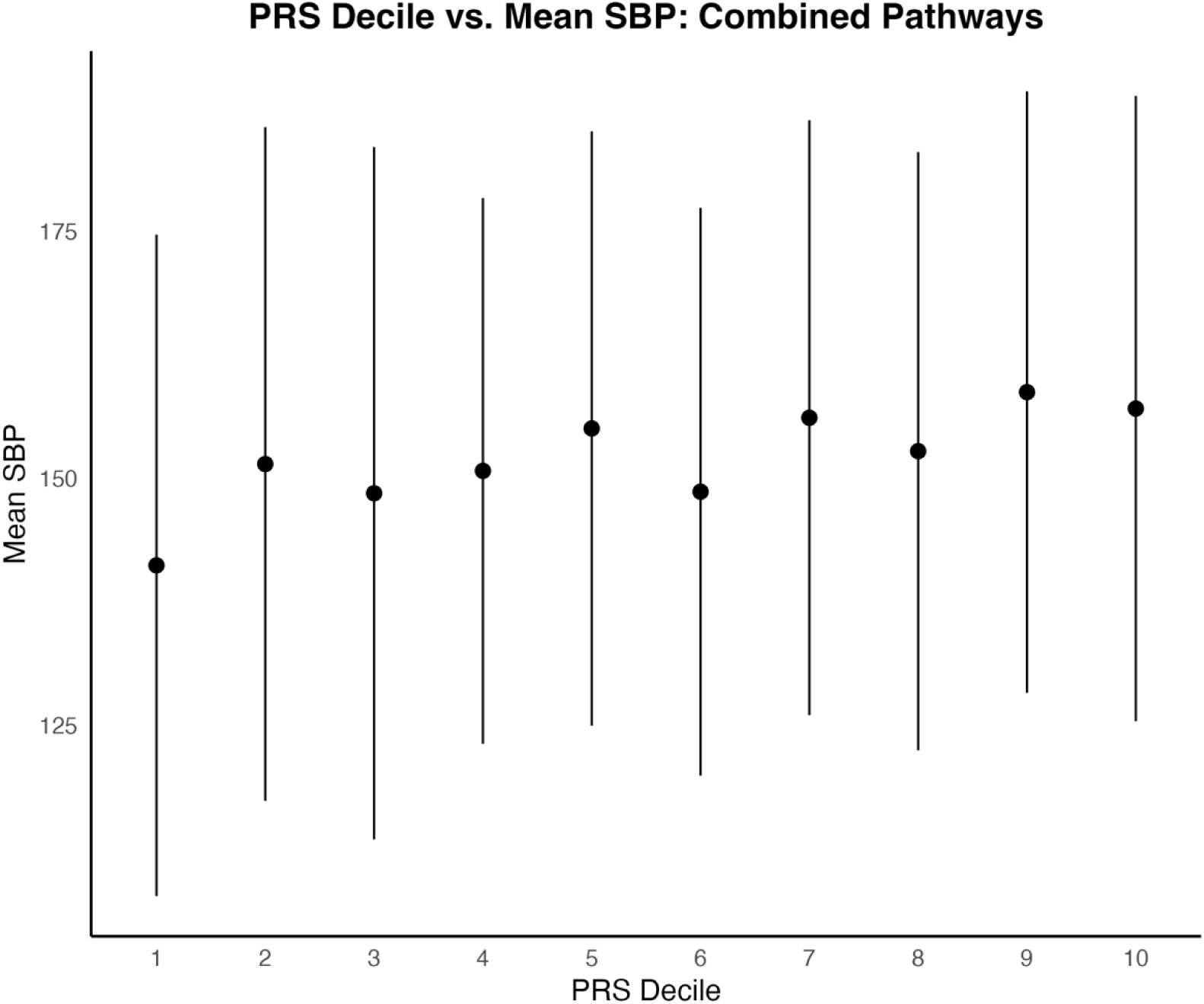
Distribution of mean diastolic blood pressures across PRS Deciles. Standard error bars of +/− one standard deviation are included.

**Figure 1b.**
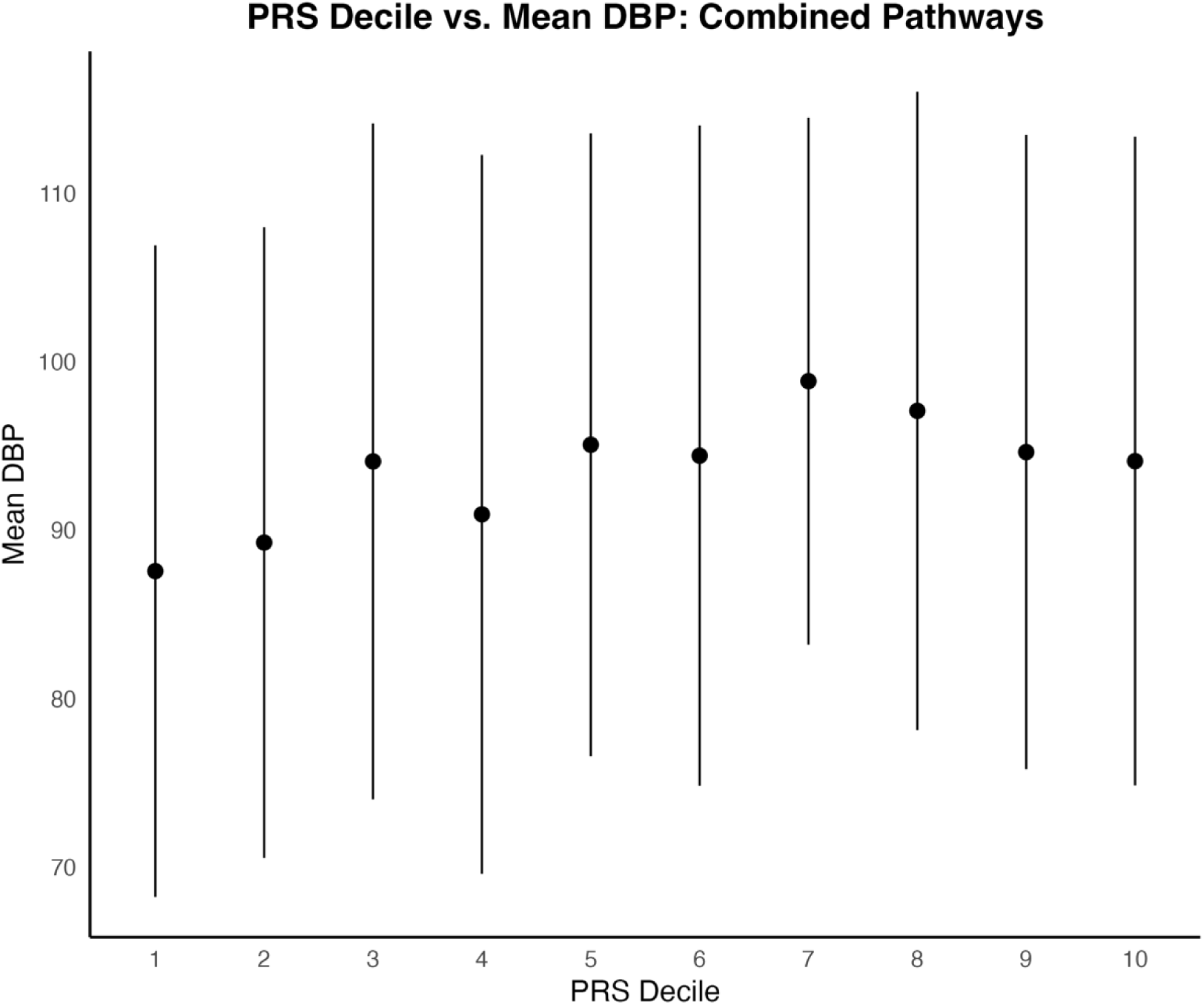
Distribution of mean diastolic blood pressures across PRS Deciles. Standard error bars of +/− one standard deviation are included.

**Figure 2a.**
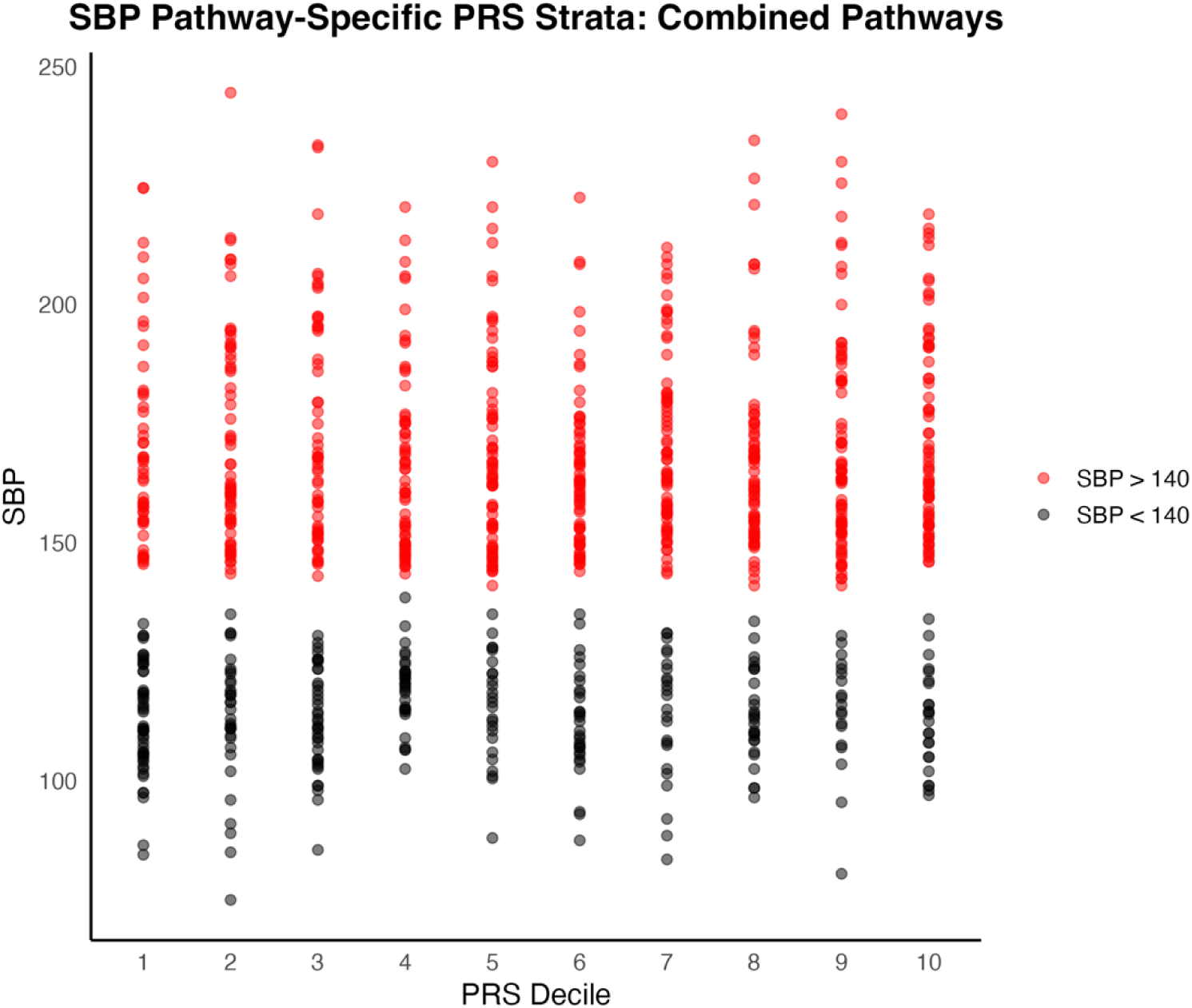
Distribution of individual SBP measures by PRS Decile. Samples are colored red for SBP greater than or equal to 140 mmHg; samples with SBP less than 140 mmHg are colored black.

**Figure 2b.**
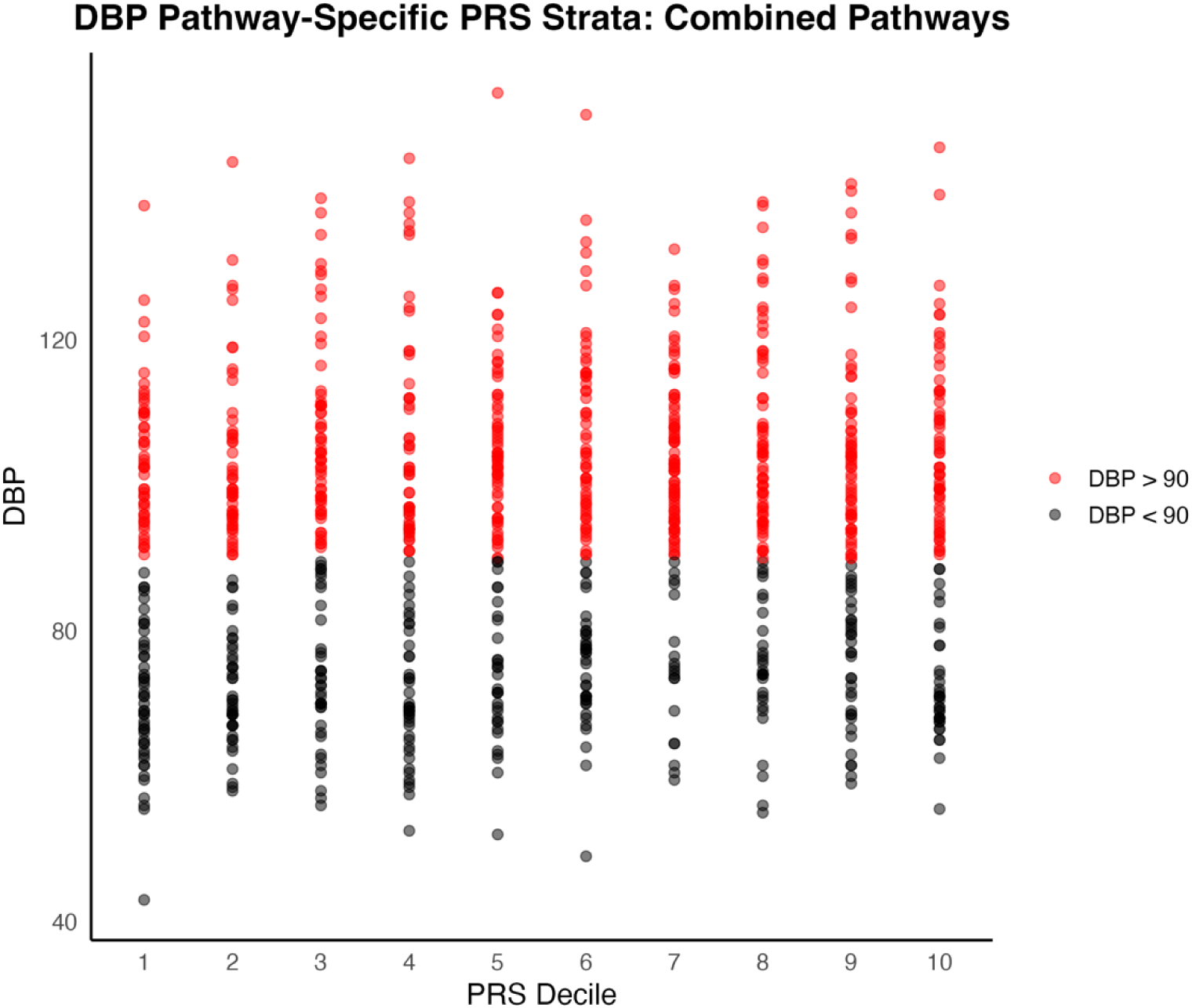
Distribution of individual DBP measures by PRS Decile. Samples are colored red for DBP greater than or equal to 90 mmHg; black for DBP less than 90 mmHg.

For DBP, 1,163 independent SNPs remained after clumping. The optimal *p-value* threshold was found to be 0.025, yielding a list of 70 SNPs for analysis, corresponding to 61 unique genes. The incremental increase in R² due to PRS for DBP was 0.0046 (1.4%, p = 0.012).

For DBP, comparing the first and last PRS deciles also yielded a significant distinction, albeit with a smaller effect size (t = −2.35, p = 0.0198, Cohen’s d = 0.34). Mean DBP in the first PRS decile was 87.1 mmHg (SD = 19.2), while mean in the last decile was 93.4 mmHg (SD = 19.6) (Figures 1b and 2b).

To assess predictive ability of each PRS for hypertension in a clinical context, we used the area under the receiver operating characteristic curve (AUC). Hypertensive patients were defined as those with systolic or diastolic blood pressures greater than 140 mmHg and 90 mmHg, respectively, or on anti-hypertension medications. The combined pathways PRS was not strongly predictive of clinical hypertension for either SBP (AUC = 0.596) or DBP (AUC = 0.557) in the target cohort (Figures 3a and Figure 3b).

**Figure 3a.**
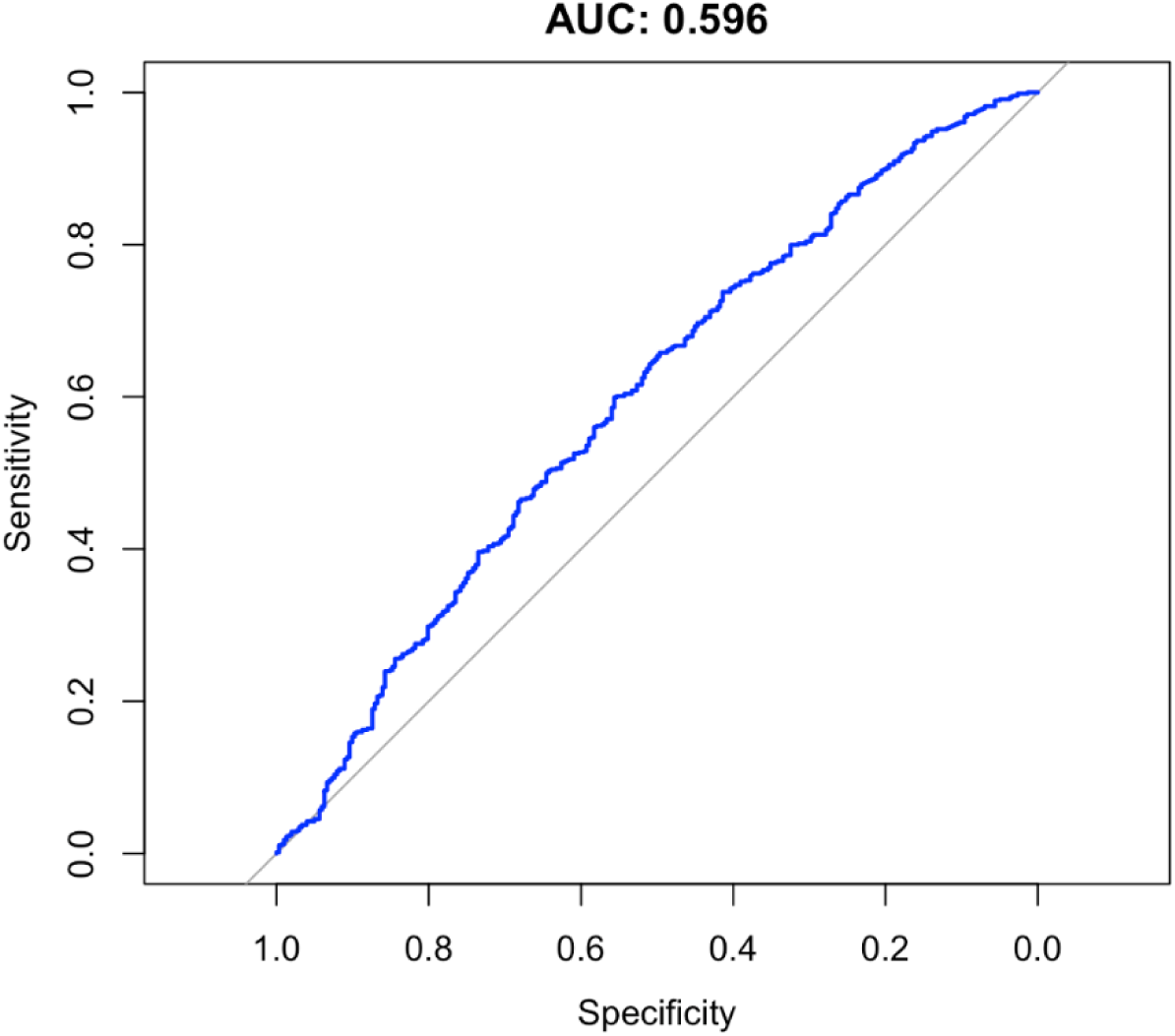
AUC for Systolic Blood Pressure Polygenic Risk Scores.

**Figure 3b.**
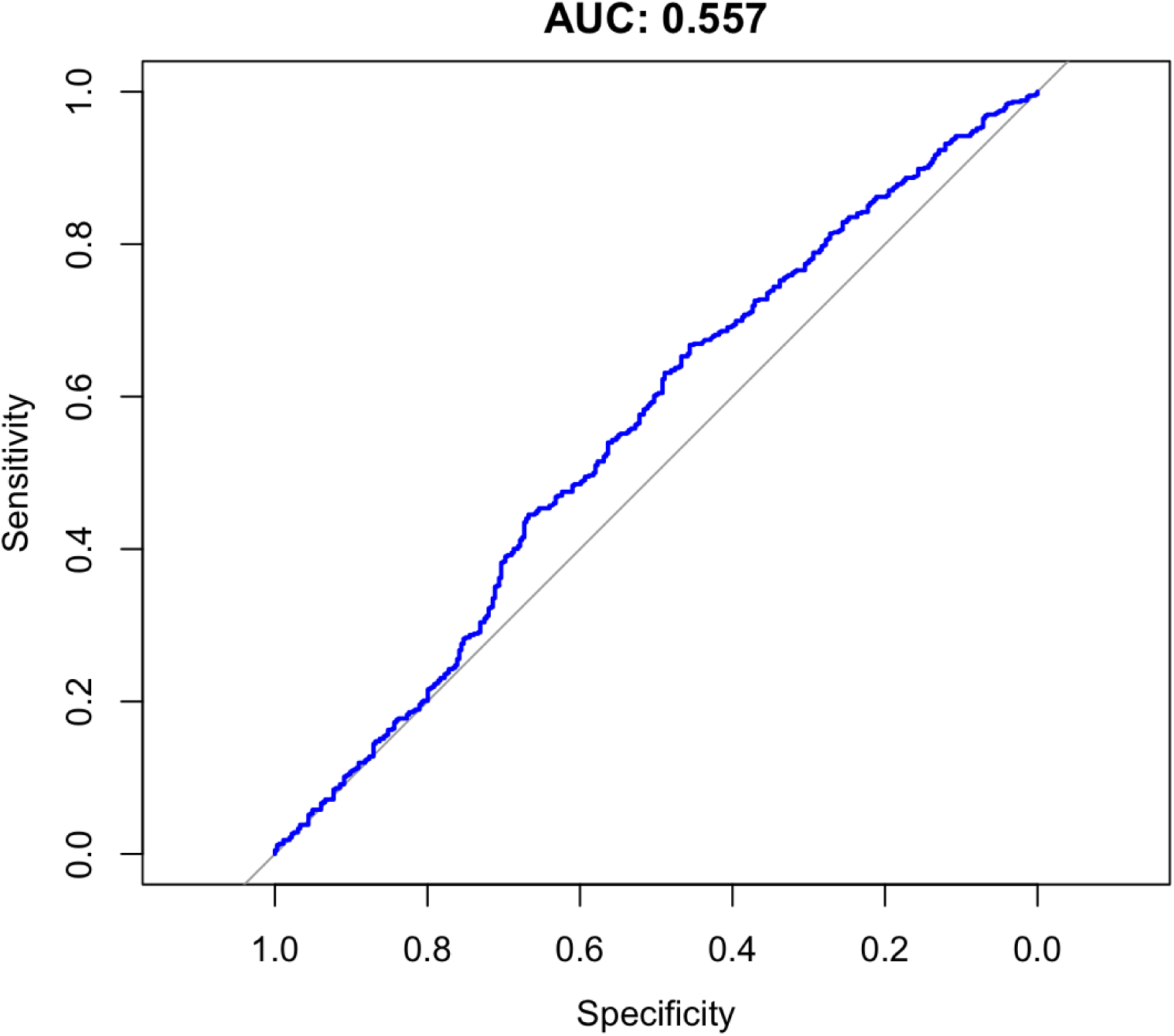
AUC for Diastolic Blood Pressure Polygenic Risk Scores.

In the MAP analysis, 1,158 independent SNPs remained after clumping. The optimal p-value threshold was found to be 0.05, yielding 109 SNPs for analysis, corresponding to 92 unique genes. The incremental increase in R² due to PRS for MAP was 0.0031 (1.1% increase, p = 0.044). When comparing the first and last deciles of PRS scores for MAP, the combined pathways PRS showed a significant distinction with a moderate effect size (t = −3.17, p = 0.0018, Cohen’s d = 0.46).

For pulse pressure, 1,149 independent SNPs remained after clumping. The optimal p-value threshold was found to be 0.05, yielding 108 SNPs for analysis, corresponding to 89 unique genes. A statistically significant increase in R² due to PRS was not observed in the pulse pressure analysis (R² = 0.0033, 5.6% increase, p = 0.066). When comparing the first and last deciles of PRS scores for pulse pressure, the combined pathways PRS did not reach statistical significance and suggested only a small effect size (t = −1.64, p = 0.103, Cohen’s d = 0.24).

### Pathway-Specific PRS

In all, 11 pathway-specific polygenic risk scores were calculated for each of four blood pressure metrics. Due to overlapping SNPs and shared genetic architecture, there was moderate correlation between PRS for many of the pathways. For example, the vascular smooth muscle contraction pathway and cGMP-PKG pathway scores for DBP had a correlation coefficient of 0.64. These pathways share 70 common genes, leading to a stronger correlation between PRS. Conversely, the renin-angiotensin system pathway shares no common genes with the cAMP signaling, MAPK signaling, and cardiac muscle contraction pathways, leading to correlation coefficients of 0.00, −0.02, and −0.01, respectively. [See supplementary material for more information on shared genes and correlation among pathways] (Figures 4a and 4b).

**Figure 4a.**
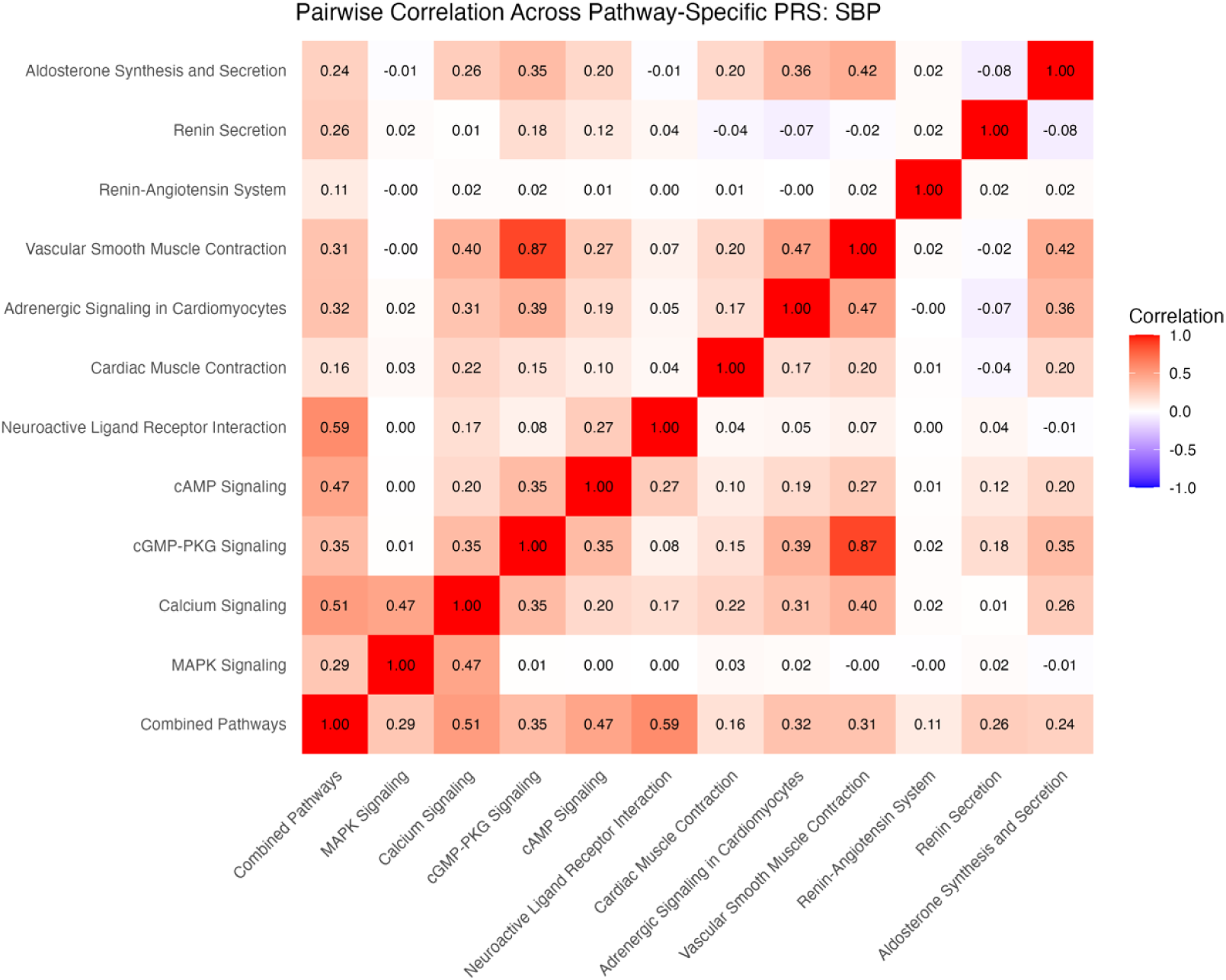
Correlations among pathway-specific PRS for SBP in discovery cohort. Correlations were calculated based on R^2^ values.

**Figure 4b.**
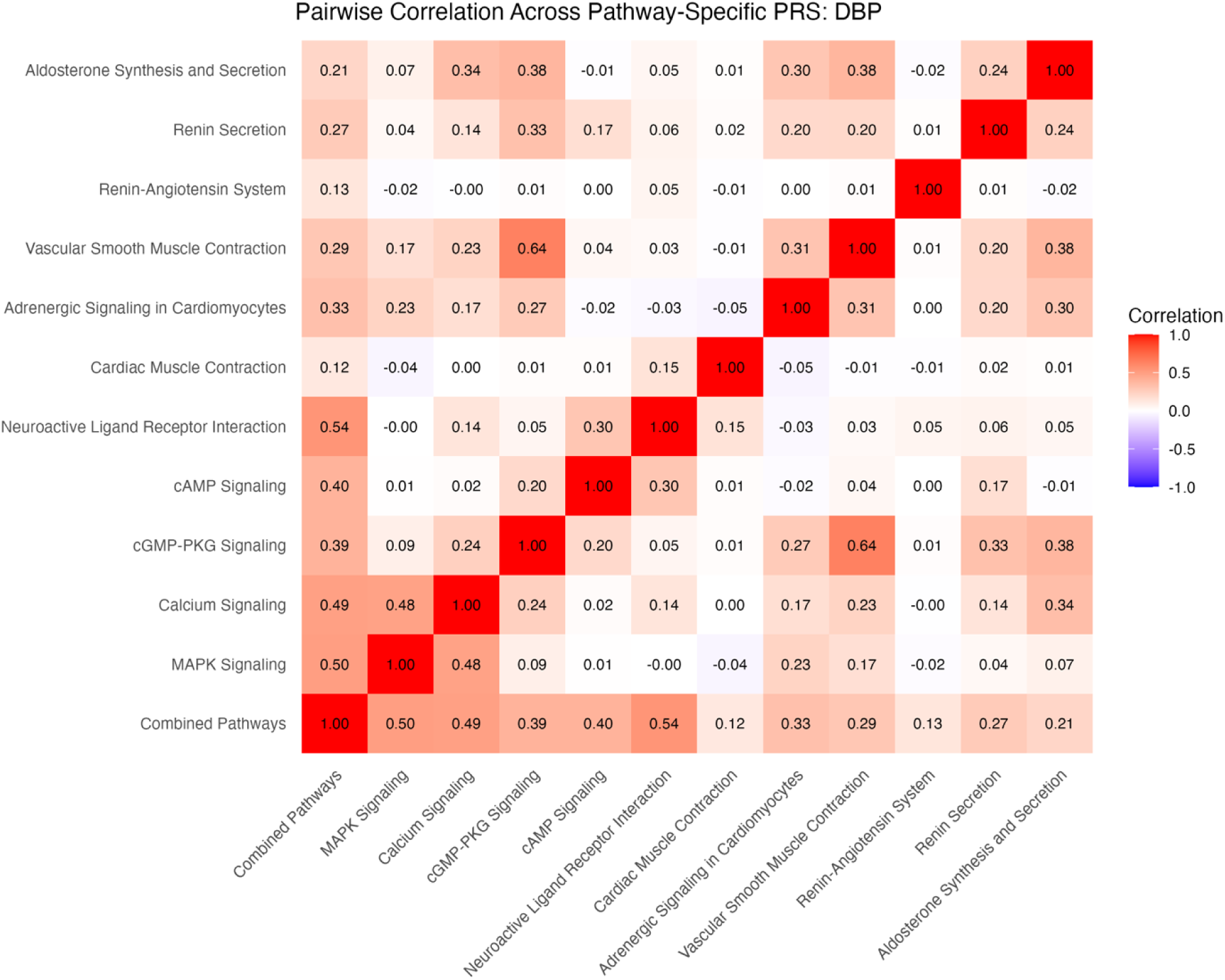
Correlations among pathway-specific PRS for DBP in discovery cohort. Correlations were calculated based on R^2^ values.

For DBP, the only pathway-specific PRS to produce a statistically significant incremental R^2^ due to PRS was the cAMP pathway (incremental R^2^ = 0.0032, 1.0% increase, *p* = 0.036.) Comparing the first and last decile of PRS scores for DBP, three pathways were able to best distinguish between high and low diastolic pressure, all with comparable effect sizes: MAPK Signaling Pathway (t-statistic = −2.78, *p* = 0.0060, Cohen’s d = 0.40), cAMP signaling pathway (t-statistic = −2.83, *p* = 0.0050, Cohen’s d = 0.41), and Adrenergic Signaling in Cardiomyocytes (t-statistic = −2.68, *p* = 0.0080, Cohen’s d = 0.39).

None of the pathway-specific PRS for SBP produced a significant increase in incremental R^2^. Also, no individual pathway-specific PRS predicted high systolic blood pressure when comparing first and last PRS deciles.

Pathway-specific PRS were also calculated for pulse pressure and mean arterial pressure. Only the Aldosterone Synthesis and Secretion pathway PRS was significantly associated with increased pulse pressure when comparing first and last PRS deciles (t-statistic=−2.77, *p* = 0.0062, Cohen’s d = 0.40). For mean arterial pressure, the Adrenergic signaling in cardiomyocytes pathway was significantly associated with increased mean arterial pressure when comparing first and last PRS deciles (t-statistic = −2.50, *p* = 0.013, Cohen’s d = 0.36).

## Discussion

In this study, a list of 1,149 unique genes were compiled from 11 different anti-hypertensive therapy-related pathways and used to calculate PRS for different blood pressure metrics in a moderate-sized non-admixed West African cohort. In a linear regression analysis, these PRS were predictive of both elevated systolic and diastolic blood pressures. However, PRS analyses specific to individual pathways were not consistently predictive of blood pressure metrics.

Multiple groups have attempted to predict phenotypes ranging from quantitative systolic and diastolic blood pressure, pulse pressure, and presence of hypertension with successful demonstration that individuals with more blood pressure-associated genetic variants of interest are more likely to have higher blood pressure.^4,24,25^ While several approaches to accomplish PRS analyses have been developed since the original reports of PRS, it is only recently that pathway-associated PRS methods have been used to study blood pressure.^3,26^ Pathway-specific PRS can be considered a form of restricted and functionally informed PRS, where the functional annotation is derived from pathways targeted by therapeutic agents for the disorder. It offers unique utility in the study of hypertension because there are multiple therapeutics, for example, angiotensin-converting-enzyme inhibitors (ACEI), angiotensin II receptor blockers (ARB), and calcium channel blockers (CCB), which mediate their effects through well-defined pathways. These helped inform the pathways we chose to study, as they all play a role in one or multiple known mechanisms of action of the above medications.

### Calcium signaling, cardiomyocyte and vascular smooth muscle contraction pathways

In this study, none of the individual pathway-specific PRS related to calcium signaling and muscle contraction were predictive of either systolic or diastolic blood pressure. Calcium signaling, through its downstream effects on various enzymes including phospholipase C and ryanodine receptors, mediates the contractility of both vascular and cardiac myocytes via the coordinated release of additional ionic calcium from the endoplasmic and sarcoplasmic reticula, respectively.^27^ This action is the primary target of calcium channel blocking agents e.g. amlodipine as they limit the re-entry of calcium into cells via the L-type calcium channel, thereby limiting the overall concentration of calcium in these tissues and reducing contractility.^27^ Angiotensin receptor blockers also contribute to the reduction of vascular smooth muscle tone by blocking angiotensin II at the AT1 receptor, interfering with phospholipase C, inositol 1,4,5-trisphosphate and diacylglycerol (PLC-IP3-DAG) signaling, and inhibiting cellular proliferation via mitofusin-2.^28,29^ A previous study showed that pathway-specific PRS based on both the cardiac muscle contraction and calcium signaling pathways were predictive of hypertension among European ancestry individuals.^30^ However, in the study cohort, the genetic variants within these pathways may not have strong enough individual effects to be predictive in a polygenic risk score.

### Neuroactive ligand receptor interaction, cardiomyocyte adrenergic and MAPK signaling pathways

In this study, the cAMP pathway PRS was predictive of elevated DBP. When comparing first and last deciles of PRS, both the cAMP and adrenergic signaling pathway PRS could reliably distinguish between high and low DBP. Additionally, the first decile of the adrenergic signaling pathway PRS was predictive of elevated mean arterial pressure when compared to the last decile. We found the first PRS decile of the MAPK pathway to be predictive of elevated DBP but not SBP compared to the last PRS decile.

Cardiomyocytes are subject to considerable sympathetic autonomic signaling by neuroactive ligands such as epinephrine and norepinephrine through both beta- and alpha-adrenergic receptors. Beta-adrenergic receptors stimulate cAMP production and downstream calcium release, ultimately resulting in stronger and faster contraction of the heart.^27^ Alpha-adrenergic receptors are similarly responsive to hormonal signaling through the PLC-IP3-DAG pathway. This mechanism is targeted by both ARB and CCB at the intracellular level as previously described, and by beta blocking agents (BB) extracellularly at neuronal-myocardial synapses.^27^ ^28^

Notably, CCB also inhibits the classical mitogen-activated protein kinase (MAPK) signaling pathway through the same mechanism. The MAPK signaling pathway is highly diverse, regulating many cellular functions from cell growth to tissue differentiation, and is sensitive to the downstream effects of reduced calcium signaling through its relation to the PLC-IP3-DAG pathway.^31^ Previous studies have linked multiple genes in the MAPK pathway to hypertension.^32,33^ and beta-adrenergic receptor polymorphisms to blood pressure variability and alterations in cardiac output,^34^ which is in line with our results.

### Renin-angiotensin system, renin, and aldosterone secretion pathways

The pathway-specific PRS related to the Renin-Angiotensin-Aldosterone system were not predictive of either systolic or diastolic blood pressures on their own. However, the aldosterone synthesis and secretion pathway PRS was significantly associated with increased pulse pressure when comparing first and last PRS deciles (t-statistic=−2.78, *p* = 0.0062).

Alongside hormonal signaling from the central nervous system, much of blood pressure determination comes from precise renal sensing and regulation.^35^ In response to low sodium, low blood pressure, or beta agonism, renin is released from the juxtaglomerular apparatus causing the downstream release and activation of angiotensin I and II, and aldosterone (RAAS).^35^ The resultant vasoconstriction and sodium salt/water retention increases blood pressure, though these pathways are frequently dysregulated in individuals with hypertension secondary to overactivation of the RAAS system and faulty compensatory reactions in the CNS.^36^ ACEI and ARB are frequently chosen first-line agents for managing essential hypertension because of their efficacy against the actions of renin and aldosterone, primary mediators of blood pressure.^28,37^ It has been shown that higher plasma aldosterone concentrations are correlated with greater pulse pressure, suggesting that elevated aldosterone levels may contribute to reduced arterial elasticity and increased pulse pressure.^38–40^ Similarly, a renin-angiotensin system pathway PRS has previously been shown to predict hypertension.^30^ However, this is notably the first instance where an aldosterone synthesis and secretion pathway PRS has been linked to increased pulse pressure.

None of the individual pathway PRS were independently significantly predictive of SBP. However, when the genes from these pathways were combined to generate one PRS, the combined PRS could reliably predict elevated SBP.

The chief strength of the pathway-specific approach is that it allows better biological interpretability of PRS. Specifically, by limiting the PRS gene sets to pathways targeted by common first-line anti-hypertensive medications, we gain better insight into clinically relevant genetic effects on blood pressure. However, this was complicated by poor predictability in our study which may have several explanations. Most notably, compared to large-scale biobank samples, we were limited by the size of our discovery (2295 samples), validation (1614 samples), and target cohorts (966 samples). PRS analyses rely upon large sample sizes as the effects of single nucleotide polymorphisms are frequently very minor, and their impacts are only seen on a very large scale. Similarly, hypertension is a complex multifactorial disease with genetic, epigenetic, and environmental influences. The genetic architecture of hypertension is highly complex and involves multiple genes with small individual effects. This complexity can obscure the predictive power of PRS, especially when considering the polygenic nature of blood pressure regulation.^41–43^

There exists an interplay between genetic and environmental factors in the development of hypertension, with some evidence that individuals genetically predisposed to high blood pressure may be more vulnerable to common modifiable risk factors.^44^ However, due to the cross sectional nature of this study, we could not account for the myriad modifiable risk factors that have been associated with hypertension risk.^42,45–47^ Additionally, there can be substantial intra-individual variability in blood pressure measurements. A 2020 meta-analysis of ambulatory blood pressure reported 95% limits of agreement for daytime systolic BP ranging from −16.7 to 18.4 mmHg, which limits interpretability of subtle changes.^48^ This variability can help explain the large standard deviations in both systolic and diastolic blood pressures observed among PRS deciles.

Notably, some of our genotyping was carried out with the Illumina Human Exome BeadChip v1.0 as described above. As this array is exome-wide as opposed to genome-wide, we do not account for the contributions of non-coding genetic information which is incompletely addressed by imputation. Additionally, this study is not intended to compare or evaluate different methods of PRS computation. We used the basic formula for PRS calculation as implemented in the software PLINK ^23^ with the understanding that other methods including PRSice, PRSice-2, and PRS-CSx may yield different results and perhaps offer greater utility in studies where a defined list of pathways was not already pre-determined.

## Conclusion

Anti-hypertensive medications exert their effects through well-defined genetic pathways. We have shown that a combined PRS constructed from the genes in these pathways are predictive of both systolic and diastolic blood pressures in a West African specific cohort. However, relatively poor predictability by pathway-specific PRS supports a broader influence of genetic, environmental and epigenetic factors contributing to hypertension that cannot be captured by pathway-specific PRS alone.

## Data Availability

All data produced in the present study are available upon reasonable request to the authors.

## Acknowledgements

The authors acknowledge the assistance of the research staff and participants in Ibadan and Igbo-Ora, Oyo State, Nigeria.

## Author Contributions

The study was initiated, conceptualized and designed by BT, GB and VR. AAA and BT contributed to the data collection and management. Data analysis was carried out by GB, VR and BT. GB, VR, TC, AC, NH, SNA, AAA and BT contributed to interpretation of results. The first draft of the manuscript was written by GB with input from VR and BT. GB, VR, TC, YZ, AC, SH, NH, SNA, AAA and BT reviewed and edited the manuscript. All the authors read and approved the manuscript.

